# Ventilation procedures to minimize the airborne transmission of viruses at schools

**DOI:** 10.1101/2021.03.23.21254179

**Authors:** L. Stabile, A. Pacitto, A. Mikszewski, L. Morawska, G. Buonanno

## Abstract

Reducing the transmission of SARS-CoV-2 through indoor air is the key challenge of the COVID-19 pandemic. Crowded indoor environments, such as schools, represent possible hotspots for virus transmission since the basic non-pharmaceutical mitigation measures applied so far (e.g. social distancing) do not eliminate the airborne transmission mode. There is widespread consensus that improved ventilation is needed to minimize the transmission potential of airborne viruses in schools, whether through mechanical systems or *ad-hoc* manual airing procedures in naturally ventilated buildings. However, there remains significant uncertainty surrounding exactly what ventilation rates are required, and how to best achieve these targets with limited time and resources. This paper uses a mass balance approach to quantify the ability of both mechanical ventilation and *ad-hoc* airing procedures to mitigate airborne transmission risk in the classroom environment. For naturally-ventilated classrooms, we propose a novel feedback control strategy using CO_2_ concentrations to continuously monitor and adjust the airing procedure. Our case studies show how such procedures can be applied in the real world to support the reopening of schools during the pandemic. Our results also show the inadequacy of relying on absolute CO_2_ concentration thresholds as the sole indicator of airborne transmission risk.

## 1 Introduction

The COVID-19 pandemic caused by the novel SARS-CoV-2 virus has put indoor environments in the spotlight since they are where virus transmission predominately occurs [1–4]. Indeed, insufficient ventilation in highly crowded environments such as restaurants, schools, and gyms does not allow proper dilution of virus-laden respiratory particles emitted by infected subjects, leading to a high percentage of secondary infections amongst exposed susceptibles [2,5–8]. To this end, governments worldwide have imposed temporary shutdowns of most indoor environments, including schools [9– 14], being in the difficult role of deciding whether to prioritize the right to education or to health. After the first pandemic wave (early 2020), guidelines for reopening schools were prepared and adopted in view of opening the schools in the late (northern hemisphere) summer, but they mainly relied upon promoting personal behaviors and basic non-pharmaceutical mitigation measures (i.e. social distancing, hand washing hand, wearing masks) that address close contact transmission [15], which is a minor route of transmission in indoor environments if a social distance in guaranteed [16,17]. The limited effect of such measures was confirmed by a resurgence of the virus in late 2020 that caused schools to close once more in many countries worldwide [18,19] (en.unesco.org/covid19/educationresponse). Thus, in order to open schools safely at the time of pandemics, airborne transmission related to the small airborne respiratory particles (droplet nuclei) [15] needs to be taken into account since it is potentially the dominant mode of transmission of numerous respiratory infections, including SARS-CoV-2 [3,20–23]; therefore, while waiting for the vaccination campaign to be completed, a suitable solution to minimize the virus transmission potential in schools is providing *ad-hoc* ventilation able to lower the virus concentration indoors [6,8,24,25].

The provision of a proper ventilation rate certainly cannot be taken for granted since most of the schools worldwide rely upon natural ventilation and manual airing (e.g. 86% of the European school buildings investigated within the SINPHONIE project [26,27]). For such schools, a potential approach to monitor and minimize the virus spread in indoor environments could be the use of a proxy providing real-time information on the virus concentration indoors then suggesting to apply manual ventilation procedures accordingly. Exhaled CO_2_ has been proposed as a possible proxy for virus transmission indoors as it is a commonly used indicator of the ventilation rate and, more generally, indoor air quality [28–30]. While in principle exhaled CO_2_ could be a good proxy for indoor-generated gaseous pollutants (e.g. VOCs, radon) [31], it cannot predict behaviors and dynamics of virus-laden particles which are affected by phenomena typical of all airborne particles such as deposition, and filtration (if any) in addition to virus inactivation. As such, the best application of exhaled CO_2_ is estimating the air exchange rate of confined spaces [32,33]. Indeed, if the particle deposition rate and virus inactivation rate are known, the indoor virus concentration is just affected by the air exchange rate; with this in mind, exhaled CO_2_ can predict the virus spreading in indoor environments and CO_2_ sensors can represent a marker of the corresponding infection risk [24,29,34,35]. Nonetheless, at this stage of the scientific debate, the question is not just demonstrating the qualitative association between ventilation (or CO_2_ levels) in buildings and the transmission of infectious diseases [3,6,8,24,28,36,37], but quantifying and guaranteeing the required ventilation in highly crowded environments (e.g. schools) to reduce the spread of infectious diseases via airborne route whether mechanical ventilation systems are installed or not.

In the present paper we evaluated the required air exchange rates for mechanically-ventilated schools and adequate airing procedures for naturally-ventilated schools to reduce the transmission potential of a respiratory virus (expressed as reproduction number) through the airborne route of transmission. Moreover, a suitable feedback control strategy, based on the continuous measurement of the indoor exhaled CO_2_ concentration, was proposed to monitor that an acceptable individual risk of infection is continuously maintained even in schools not equipped with mechanical ventilation systems. To this end, simulations based on virus and exhaled CO_2_ mass balance equations considering typical school scenarios were performed.

## 2 Materials and methods

The required air exchange rates and the adequate airing procedures to maintain an acceptable level of the virus transmission risk were calculated adopting the virus and CO_2_ mass balance equations (described in section 2.1 and 2.2) under the simplified hypothesis that they are both instantaneously and evenly distributed in the confined space under investigation (box-model). Here particle deposition and virus inactivation phenomena were taken into account and dynamic scenarios (described in section 2.3) have been simulated within the 5-hour school-day. Two different viruses, characterized by extremely different emission rates (i.e. different viral loads and infectious doses) [38], were considered: SARS-CoV-2 and seasonal influenza. The study involves infected people breathing and/or speaking whereas severely symptomatic persons frequently coughing or sneezing were not included in the scenarios. The simulations were performed under the hypothesis that the students are adequately spaced so that ballistic deposition of large respiratory particles (> 100 µm) onto mucous membranes is considered negligible [15]; thus, virus transmission results solely from the Inhalation of airborne particles (i.e. airborne transmission).

### 2.1 Evaluation of the virus transmission potential

The virus transmission potential due to the airborne route was assessed in terms of event reproduction number (R_event_) which is the expected number of new infections arising from a single infectious individual at a specific event [39] (e.g. a single school day). In particular, the R_event_ was evaluated adopting the approach proposed and applied in previous papers [5,6,40]; involving six successive steps: (i) the quanta emission rate, (ii) the exposure to quanta concentration in the microenvironment, (iii) the dose of quanta received by exposed susceptible subjects, (iv) the probability of infection on the basis of a dose-response model, (v) the individual risk of the exposed person, and, finally, (vi) the event reproduction number. The above-mentioned “quanta” is a measure to quantify the virus emission or concentration, it is defined as the infectious dose for 63% of susceptibles by inhalation of virus-laden particles. In particular, the evaluation of the quanta emission rate (ER_q_, quanta h^-1^) was described in our previous papers taking into account the viral load, infectious dose, respiratory activity, activity level, and particle volume concentration expelled by the infectious person [5,6,38]. The model, here not reported for the sake of brevity, provides a distribution of quanta emission rates, i.e. the probability density function of ER_q_. It represents a major step forward to properly simulate and predict infection risk in different indoor environments via airborne transmission since previous studies were performed adopting quanta emission rates obtained from rough estimates based on retrospective assessments of infectious outbreaks only at the end of an epidemic [24,41]. The predictive approach also enables stochastic analysis of infection probability that is not possible when using a point estimate obtained from a superspreading event.

The indoor quanta concentration over time, n(t,ER_q_), is evaluated, for each possible ER_q_ value, adopting the above-mentioned simplified mass balance equation as:

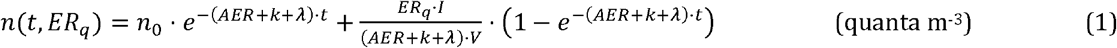

where AER (h^-1^) is the air exchange rate, k (h^-1^) is the deposition rate on surfaces, λ (h^-1^) is the viral inactivation rate, I is the number of infectious subjects, and V is the volume of the indoor environment. The dose of quanta (D_q_) received by a susceptible subject exposed to a certain quanta concentration for a certain time interval, T, can be evaluated by integrating the quanta concentration over time as:

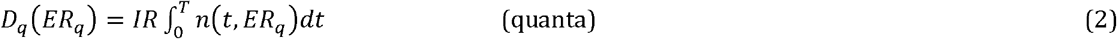

where IR is the inhalation rate of the exposed subject which is a function of the subject’s activity level and age [42,43].

The probability of infection (P_I_, %) of exposed persons (for a certain ER_q_), is evaluated on the basis of simple Poisson dose-response model [44,45] as:

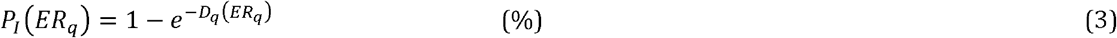

The individual risk of infection (R) of an exposed person for a given exposure scenario is then calculated integrating, over for all the possible ER_q_ values, the product between the conditional probability of the infection for each ER_q_ (P_I_(ER_q_)) and the probability of occurrence of each ER_q_ value (P_ERq_):

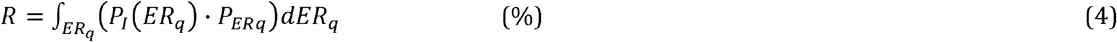

Such an individual risk R, for a given exposure scenario, represents the ratio between the number of new infections (number of cases, C) and the number of exposed susceptible individuals (S). The R_event_ (expected number of new infections, C, arising from a single infectious individual, I, at a specific event) can be obtained as the product of R and S:

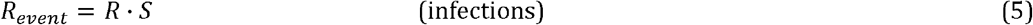

Therefore, the maximum number of susceptibles that can stay simultaneously in the confined space under investigation for an acceptable R_event_ < 1 (hereinafter referred as maximum room occupancy, MRO) is:

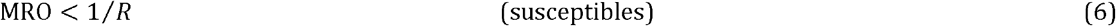

### 2.2 Evaluation of the CO2 indoor levels

To estimate the trend of indoor (exhaled) CO_2_ concentration over time (CO_2-in_) a mass balance equation was applied considering the initial indoor CO_2_ concentration (at t=0) equal to outdoor air (CO_2-out_), the mass balance equation can be simplified as [32]:

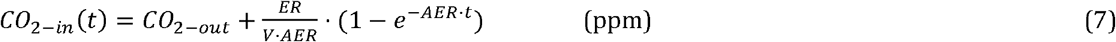

where ER represents the overall exhaled CO_2_ emission rate in the indoor environment under investigation; the emission rate per-capita are available in the scientific literature (typically expressed in L s^-1^ person^-1^) as a function of the activity level, age, and gender [46]. As mentioned above, for known and steady state emission rate and outdoor CO_2_ concentration, the indoor concentration is just affected by the air exchange rate of the room, and the AER can be back-calculated from the eq. 7 measuring continuously the indoor CO_2_ concentration (CO_2-in_): this measurement method is known as “constant injection rate method” [32,47].

### 2.3 Simulated scenarios

The individual risk of infection and the event reproduction number of a disease due to the airborne transmission route of the virus were assessed considering a high-school classroom (e.g. students aged 17-18) with a floor area of 50 m^2^ and a height of 3 m (V=150 m^3^). A crowding index suggested by the standard EN 15251 [48] on the design of the ventilation for a proper indoor air quality (2 m^2^ person^-1^) was adopted then obtaining a total number of occupants (including the teacher) of 25 persons. A total school-time of 5 hours was considered. The simulations were performed considering one infected subject (I=1), the teacher or one of the students, and 24 exposed susceptibles (S=24) hypothesizing that none of them is already immune (e.g. vaccinated). Therefore, in order to obtain a R_event_ < 1, the individual risk of infection (R) of the exposed susceptible over the 5-hour school-time should be less than 1/24, i.e. < 4.2%.

The simulations were conducted for different scenarios, i.e. combination of emitting subject and mitigation solution (if any). Two different emitting subjects were considered in the simulation: the teacher and the student. In particular, simulations were performed considering (a) the infected teacher giving lesson (i.e. speaking or loudly speaking) for one hour, in particular, the first hour of lesson was considered as it is clearly the worst exposure scenario for susceptible students attending the lesson (in fact, the latest the infected teacher enters the classroom, the shorter the exposure period of the susceptible persons), or (b) the infected student attending lessons, then just breathing, and/or speaking occasionally. The exposed susceptibles were considered performing activities in a sitting position then inhaling at IR = 0.54 m^3^ h^-1^ [42,43].

The emitting scenarios are summarized in Table 1, whereas the corresponding quanta emission rate probability distribution function for SARS-CoV-2 and seasonal influenza viruses, as a function of activity level (i.e. sitting) and respiratory activity, are summarized in Table 2 as obtained from previous papers [5,6]. The ER_q_ clearly increases for more severe respiratory activities, e.g. the median SARS-CoV-2 ER_q_ ranges from 0.575 quanta h^-1^ for oral breathing to 15.85 quanta h^-1^ for loudly speaking. Moreover, due to its higher infectious dose (i.e. RNA copies to reach a quanta), for similar activity levels and respiratory activities, the SARS-CoV-2 ER_q_ values were much higher than the seasonal influenza ones [5,38,49–51] (e.g. more than 10-fold at median value).

**Table 1.**
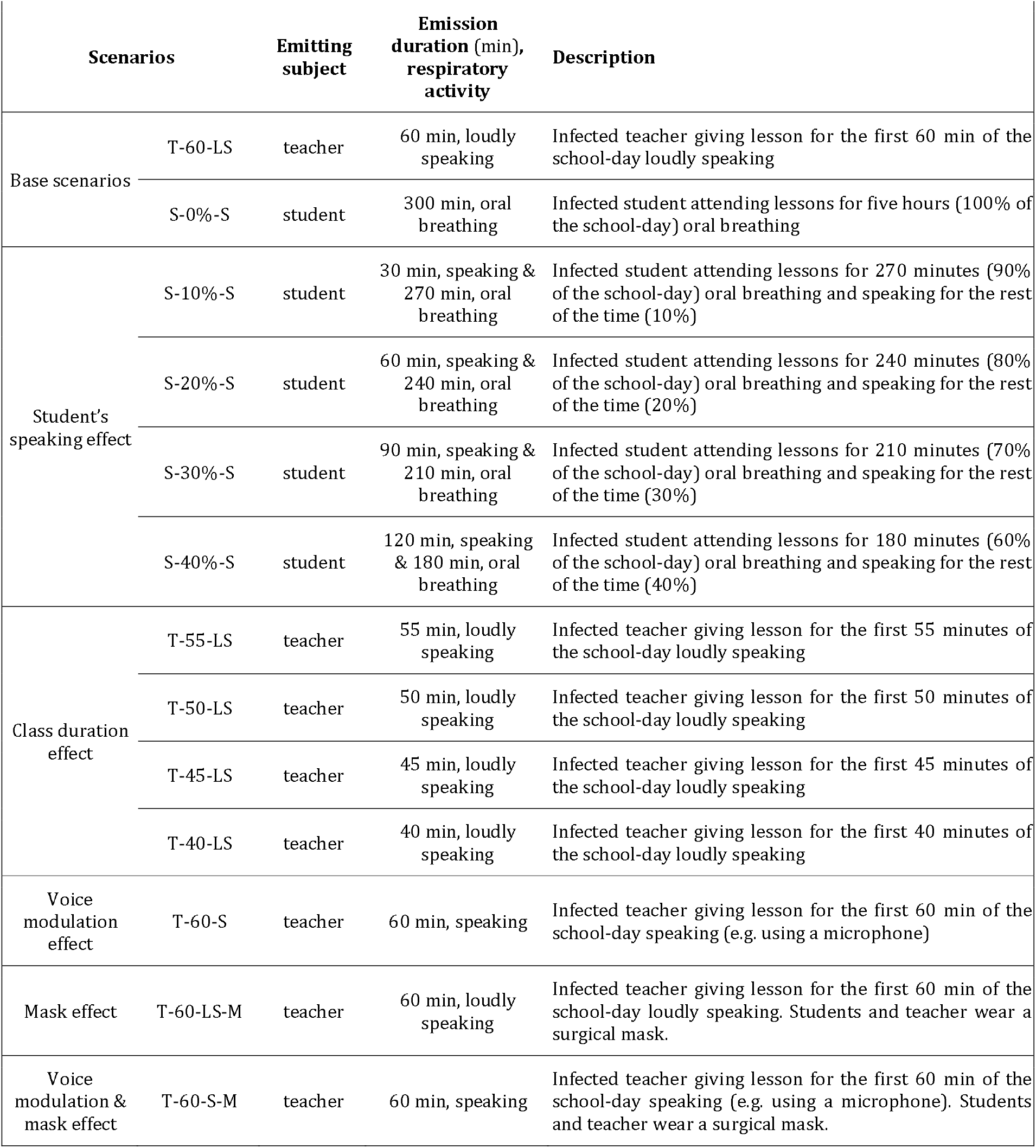
Scenarios considered to simulate the exposure to SARS-CoV-2 and seasonal influenza viruses in the classroom: emitting subjects, emission duration, and respiratory activity are summarized (whereas the same activity level, i.e. sitting, was considered for all the scenarios). Descriptions of the base scenarios and of the possible mitigation solutions are reported.

**Table 2.**
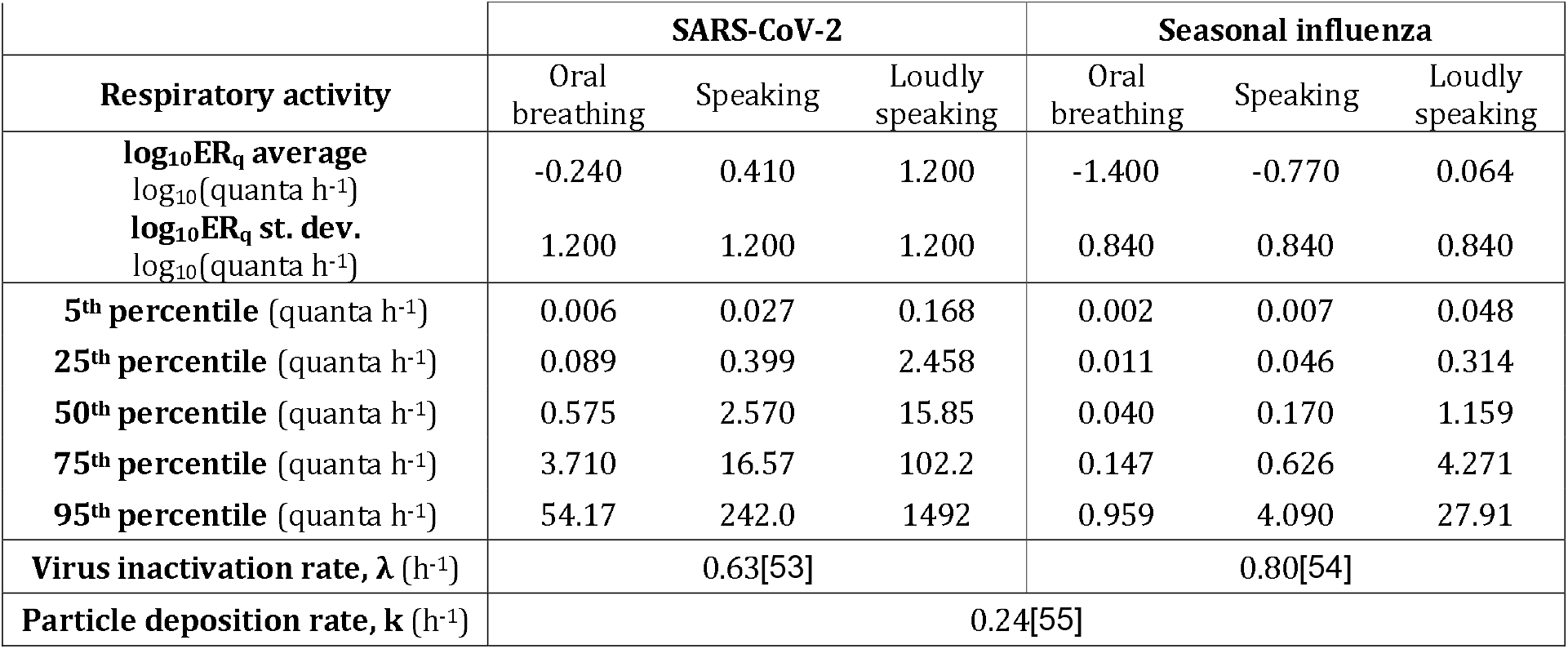
Quanta emission rate distribution (ER_q_, quanta h^-1^), expressed as log_10_ average and standard deviation values as well as 5^th^, 25^th^, 50^th^, 75^th^, and 95^th^ percentiles, for SARS-CoV-2 and seasonal influenza viruses as a function of respiratory activity. Virus inactivation rate, λ (h^-1^), and particle deposition rate, k (h^-1^) are also

Despite the base scenarios, as summarized in Table 1, the possible effects of infected student’s speaking duration (10% to 40% of the time), class duration (school hour of 55, 50, 45, or 40 min instead of 60 min), infected teacher’s voice modulation (e.g. using microphone), and wearing mask were considered in the simulations and described in detail. The effect of the mask was simulated considering an overall 40% reduction of the dose of quanta received by the susceptibles [52], to this end, in such simulations the ER_q_ values were halved.

The emission rate of exhaled CO_2_ was evaluated considering a per-capita emission rate equal to 0.0044 L s^-1^ person^-1^ as an average value between males and female teenager students (e.g. aged 17-18) with a level of physical activity of 1.3 met [46], which is the suggested level for reading, writing, typing in a sitting position at school. The overall emission rate (ER) was evaluated multiplying the per-capita emission rate by the number of student/teacher (25 person), then it resulted equal 0.110 L s^-1^. In the simulations here proposed the outdoor CO_2_ concentration (CO_2-out_) was set at 500 ppm.

### 2.4 Required air exchange rates and airing procedures

The required air exchange rate to maintain a R_event_ < 1 in mechanically-ventilated schools for the abovementioned scenarios was calculated adopting the methodology described in section 2.1 and, especially, the eq. 1-5. Quanta emission rates were selected from Table 2 on the basis of the activity of the emitting subject (Table 1), the geometry of the classroom were reported in the section 2.3, the virus inactivation rate (λ) for SARS-CoV-2 (0.63 h^-1^) [53] and seasonal influenza (0.80 h^-1^) [54] as well as the particle deposition rate (k=0.24 h^-1^) [55] were obtained from the scientific literature and are summarized in Table 2. Having set these data, the individual risk of infection and, consequently, the event reproduction number, were just affected by the air exchange rate and the airing procedure of the classroom.

Quantifying the air exchange rate for mechanical ventilation systems is straightforward, as the fresh air ventilation rate can be easily measured in most cases, and should be consistent with the original design parameters of the system (assuming proper installation and routine maintenance).

For schools not equipped with mechanical ventilation systems, which are the majority [26,27], to maintain a R_event_ < 1, *ad-hoc* manual airing procedures based on manual airing cycles [31,56], i.e. adopting periods with windows closed and open alternatively, have to be determined. Indeed, unlike mechanical ventilation systems which are able to provide constant air exchange rate, the manual airing cycles will alternate periods at low air exchange rates (with window close) and periods at higher air exchange rates (with window open), and most importantly, such air exchange rates are not known a *priori*. Thus, for naturally-ventilated schools, an air exchange rate of the manual airing procedure can be calculated a-posteriori as school-day average resulting from the airing cycles (i.e. weighted average air exchange rate):

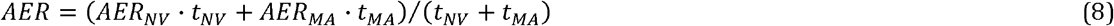

where AER_NV_ and AER_MA_ are the air exchange rates with window close (natural ventilation, NV) and window open (manual airing, MA), respectively, and t_NV_ and t_MA_ represent the total time during which the windows were kept closed and open, respectively; the sum of t_NV_ and t_MA_ clearly is the overall school time (i.e. 300 min). Since the air exchange rate is not constant all over the school day, the time at which the airing is adopted can significantly affect the quanta concentration trends. In fact, if a high quanta emission occurs when the windows are closed, the susceptibles could be exposed to high quanta concentrations then leading to a dose of quanta (and then an individual risk) larger than expected for a constant air exchange rate. In other words, for a certain exposure scenario, even when a school-day average AER provided with manual airing cycles is equal to the mechanical ventilation one, higher dose of quanta and individual risk can happen. Thus, in the case of manual airing cycles, higher average air exchange rates are needed to maintain a R_event_ < 1 with respect to classrooms equipped with mechanical ventilation systems, in particular for high but brief virus emissions. In our simulations, the manual airing cycles were applied at the end of each school-hour (instead of at the beginning of each lesson or between lessons), this just represents a constraint adopted in order to limit the number of scenarios to be simulated, nonetheless, it does not undermine the findings and the procedures we described.

Modeling air exchange rates from natural ventilation is extremely complex as leakages of the building (AER_NV_) and to the airing (AER_MA_) are strongly influenced by the airtightness of the building and of the windows, the wind conditions, the windows positioning within the classroom (single-sided vs. cross ventilation), and the window opening angle [56–59]. As an example, previous papers [31,58] performed experimental campaigns to measure the air exchange rate with window closed and opened through a CO_2_ decay method in classrooms and obtained significant variations of AER_NV_ (< 0.3 h^-1^) and AER_MA_ (up to 5 h^-1^). Summarizing, the ventilation rate via natural ventilation and manual airing is not controlled; therefore, in view of maintaining a R_event_ < 1 a feedback mechanism (the indoor CO_2_ concentration) is needed. We develop and apply this proper feedback control strategy to help optimize ad hoc airing in classrooms.

## 3 Results and discussions

### 3.1 Required air exchange rates for mechanically-ventilated schools

Figure 1 presents the trends of quanta concentration, individual risk, Maximum Room Occupancy, and indoor CO_2_ concentration for the scenarios T-60-LS (teacher giving lesson loudly speaking for the first 60 min of the school-day) and T-60-S (i.e. speaking using a microphone instead of loudly speaking) in the case of SARS-CoV-2 virus when required AERs (to maintain a R_event_ < 1) are provided through mechanical ventilation systems. In particular, for the scenario T-60-LS, as summarized in Table 3, the required AER is 9.5 h^-1^ (i.e. > 15 L s^-1^ person^-1^). The quanta concentration trend increases sharply in the first 60 min (i.e. when the virus source is still in the classroom), then exponentially decays as soon as the teacher leaves the room and goes to zero at about 90 min. The individual risk reaches the maximum permitted value (4.2%) at 90 min, then remaining constant up to the end of the school-day (300 min) as we hypothesized that no other infected people enter the classroom. Similarly, as designed, the maximum occupancy decreases to the needed value of 24 persons at the end of the school-day. The authors point out that in the scenario T-60-LS the whole dose of quanta (and then individual risk) is received by the susceptibles in roughly 90 minutes, thus, we would have designed the same air exchange rate also in the hypothesis that the infected teacher gave a lesson at the second, third or fourth hour. Due to the high (and constant) AER = 9.5 h^-1^, the CO_2_ indoor reaches the (very low) equilibrium concentration of approximately 750 ppm in about half an hour.

**Table 3.** Required constant AER (h^-1^) to maintain a R_event_ < 1 for all the scenarios investigated for SARS-CoV-2 for mechanically-ventilated classrooms.

| Scenarios | | AER ($\text{h}^{-1}$) |
| --- | --- | --- |
| Base scenarios | T-60-LS | 9.5 |
|  | S-0%-S | 0.8 |
| Student's speaking effect | S-10%-S | 1.5 |
|  | S-20%-S | 2.1 |
|  | S-30%-S | 2.8 |
|  | S-40%-S | 3.5 |
| Class duration effect | T-55-LS | 8.6 |
|  | T-50-LS | 7.8 |
|  | T-45-LS | 6.9 |
|  | T-40-LS | 6.1 |
| Voice modulation effect | T-60-S | 0.8 |
| Mask effect | T-60-LS-M | 5.8 |
| Voice modulation & mask effect | T-60-S-M | 0.2 |

**Figure 1.**
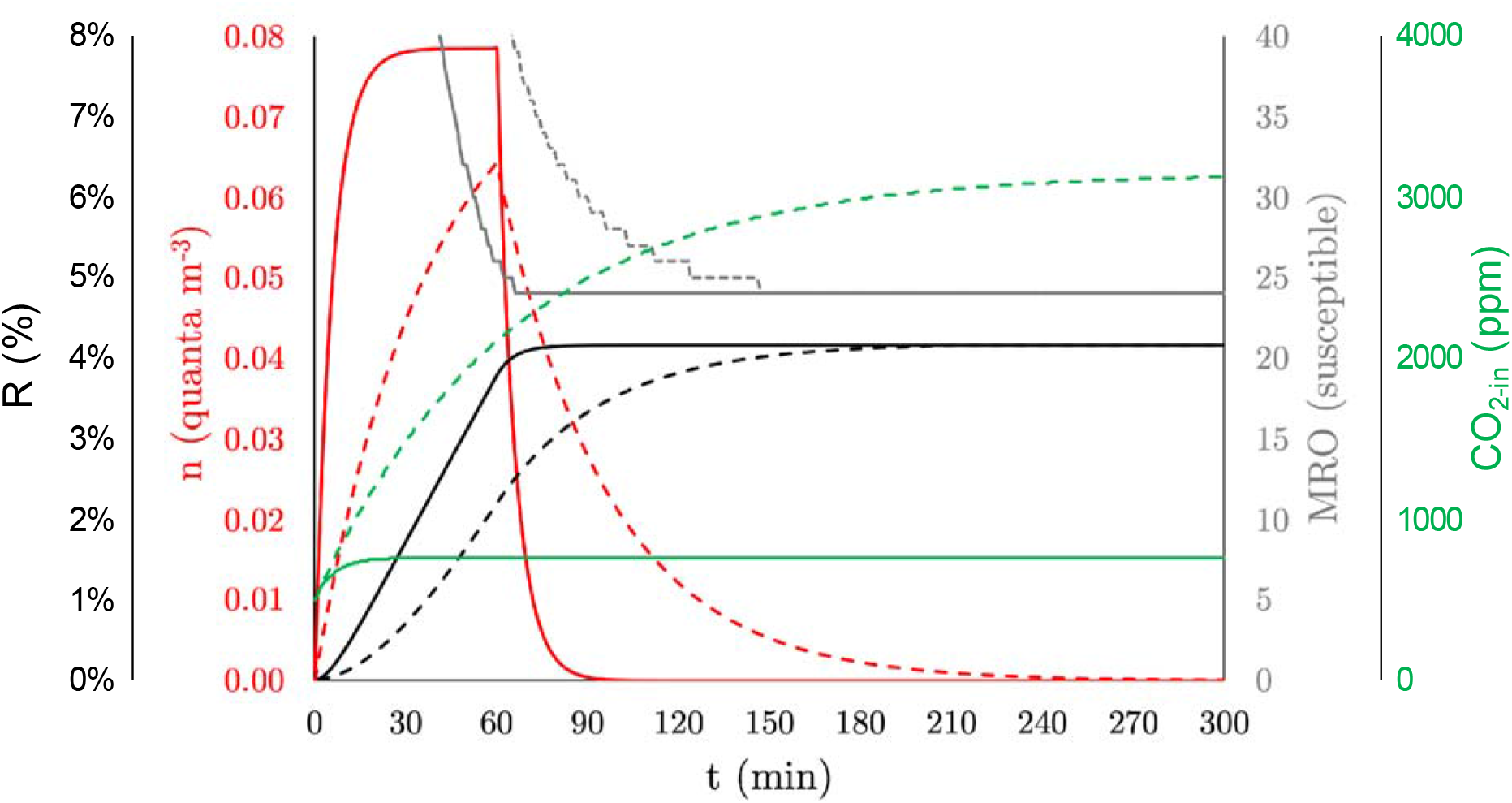
Trends of quanta concentration (n), individual risk (R), Maximum Room Occupancy (MRO), and indoor CO_2_ concentration (CO_2-in_) resulting from the simulation of the base scenarios T-60-LS (infected teacher giving lesson loudly speaking for the first 60 min of the school day, solid lines) and T-60-S (infected teacher giving lesson speaking using a microhone for the first 60 min of the school day; dotted lines) in the case of SARS-CoV-2 virus having adopted the required constant AERs to maintain a R_event_ < 1 (9.5 h^-1^ and 0.8 h^-1^ for T-60-LS and T-60-S, respectively) through a mechanical ventilation system.

For the scenario T-60-S, a much lower AER (0.8 h^-1^; i.e. 1.3 L s^-1^ person^-1^) is required to maintain a R_event_ < 1, indeed, the CO_2_ indoor concentration does not even reach an equilibrium level and continuously increases above 3000 ppm in five hours, which is well above the concentrations suggested and obtainable if EN 15251 indoor air quality standards are adopted [48].

In Table 3 the required AERs to maintain a R_event_ < 1 for all the investigated scenarios are reported for SARS-CoV-2 for mechanically-ventilated classrooms; the required AER for seasonal influenza-infected subjects is not reported since it is < 0.1 h^-1^ for all the scenarios under investigation. Thus, all the ventilation techniques are able to protect against the spreading of the seasonal influenza virus in classroom through airborne transmission. On the contrary, for SARS-CoV-2-infected subjects, the required AERs can be quite high: as mentioned above, for a teacher giving lesson for one hour the required AER is 9.5 h^-1^. Such AER can be reduced adopting shorter lessons (e.g. for 40-min lessons it can decrease down to 6.1 h^-1^) or, even more, as discussed above, keeping the voice down while speaking (e.g. using microphones, this would require just 0.8 h^-1^) and simultaneously wearing masks (then lowering the required AER down to 0.2 h^-1^). If the infected subject is a student, an AER of 0.8 h^-1^ is needed if she/he does not speak for the entire school-day, then increasing for longer speaking periods (e.g. 3.5 h^-1^ are required if she/he speaks for 40% of the school-day).

In Figure 2 the individual risk, R, of students for different exposure scenarios characterized by the presence of a SARS-CoV-2-infected teacher giving lesson for 60 min as a function of the air exchange rate provided by a mechanical ventilation system is presented. In particular, the base scenario (teacher loudly speaking) and the mitigation solutions (voice modulation and use of mask) are graphed. As expected the individual risk clearly decreases for higher AERs and, as summarized in Table 3, very high AERs are required for the teacher when loudly speaking. Such high AERs are likely not reproducible in schools without mechanical ventilation systems; indeed, in our previous papers [31,56,58,60] we have estimated that the AER, when no airing procedures are imposed, are typically lower than 1 h^-1^ and that the AER for manual airing (mainly side-ventilation) are < 5 h^-1^. Figure 2 presents the expected peak CO_2_ \concentrations (i.e. at the end of the school-day) as a function of the AERs and clearly shows that the CO_2_ level per se could be extremely misleading when not interpreted with a specific focus on infection transmission. Indeed, even when acceptable CO_2_ levels are provided (e.g. <1000 ppm), an unacceptable individual risk can occur. For high-emitting activities (i.e. loudly speaking) the mitigation solutions (e.g. the use of microphones) are more effective than the classroom ventilation itself. Furthermore, there is a transient aspect to the problem when CO_2_ concentrations start at a low level and then build up to an established acceptable level, all the while inhalation of infectious particles (droplet nuclei) may be occurring.

**Figure 2.**
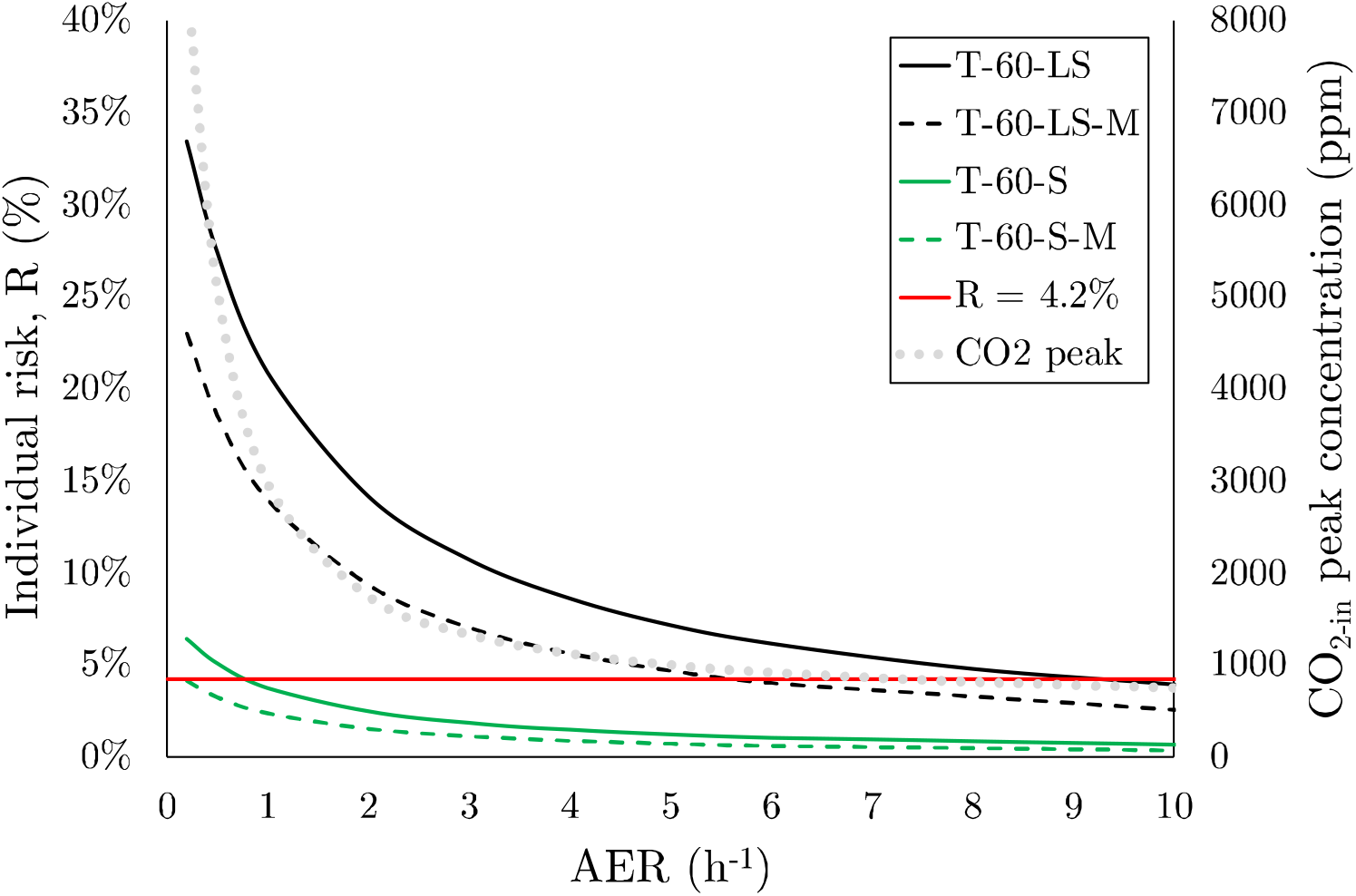
Individual risk, R (%), of students for different exposure scenarios characterized by the presence of a SARS-CoV-2 infected teacher giving lesson for 60 min as a function of the air exchange rate for mechanically-ventilated classrooms: loudly speaking (T-60-LS), speaking (T-60-S), loudly speaking and wearing mask (T-60-LS-M), speaking and wearing mask (T-60-S-M). Expected CO_2_ peak concentrations (i.e. at the end of the school-day) as a function of the AERs are also reported.

The high AERs required in some of the above-mentioned scenarios can be higher than those typically suggested by the current indoor air quality standards defined by the EN 15251 [48]. Indeed, the EN 15251 provides the AERs as a function of the category of the indoor air quality (I, II, or III) and categories of pollution from building itself (very low polluting building, low polluting building, non low polluting building). As an example, for the classroom under investigation the air exchange rates suggested by the standard would be: 6.6 h^-1^ (very low polluting building), 7.2 h^-1^ (low polluting building) and 8.4 h^-1^ (non low polluting building) for building category I, 4.6 h^-1^, 5.0 h^-1^, and 5.9 h^-1^, for building category II, and 2.6 h^-1^, 2.9 h^-1^, and 3.4 h^-1^ for building category III, respectively. Thus, for such a critical scenario, in order to maintain a R_event_ < 1 at lower air exchange rates the number of susceptibles (S) should be reduced; to this end the most effective solution is increasing the vaccination fraction of the population. As an example, in Figure 3 the R_event_ as a function of the air exchange rate (provided through a mechanical ventilation system) and of the percentage of vaccination for the T-60-LS scenario is reported. The figure clearly highlights that for such a critical scenario a percentage of vaccinated people > 60% (i.e. > 14 persons) would allow reducing the required air exchange rate to about 3 h-1, i.e. to the AER suggested by the EN 15251 for building category III which is, by the way, the category recommended by the standard for existing buildings. The graph also confirms that reopening naturally ventilated schools by allowing up to 50% attendance as adopted in several countries would not guarantee a low transmission potential of the SARS-CoV-2, at least for highly emitting infected subjects.

**Figure 3.**
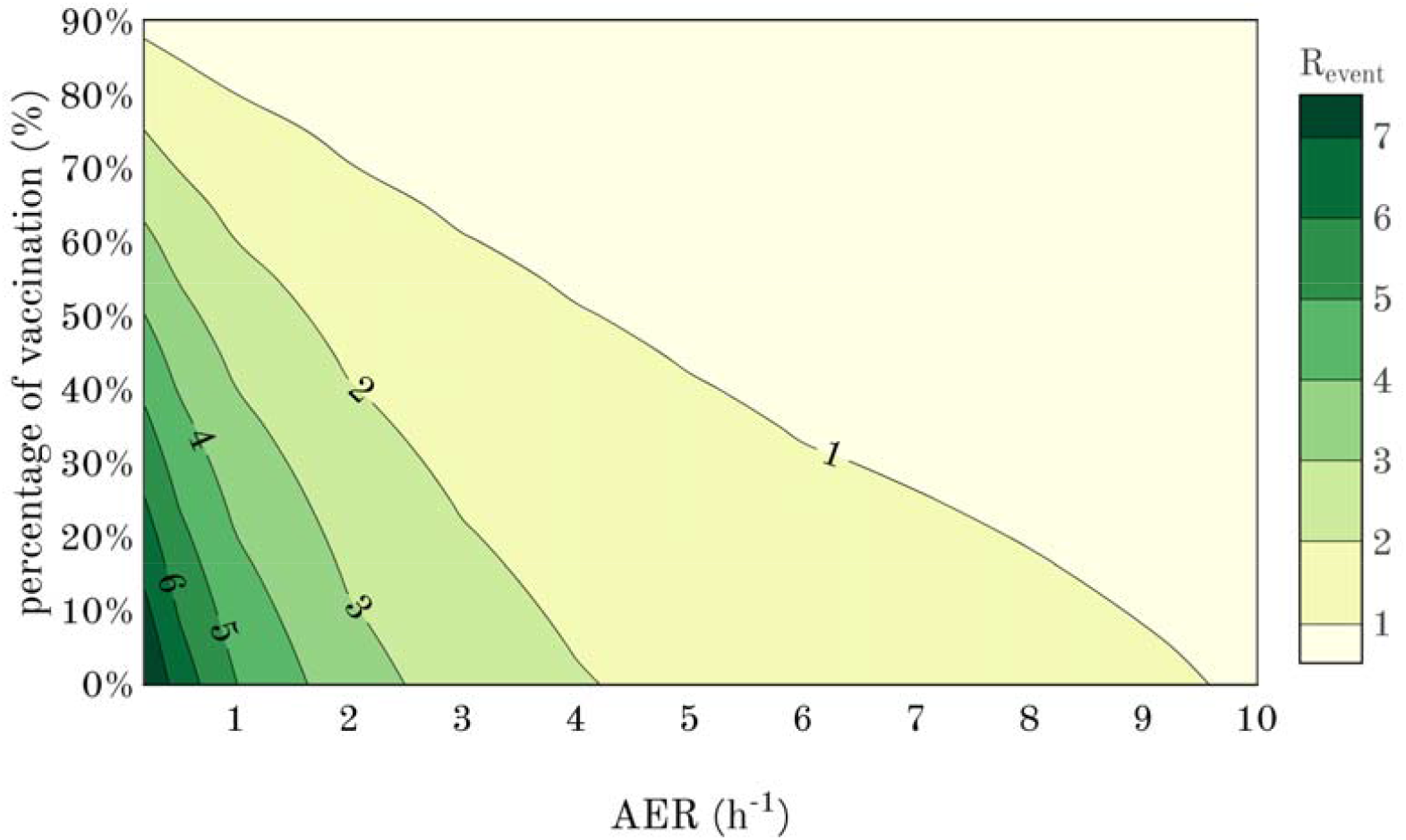
R_event_ for the T-60-LS scenario as a function of the air exchange rate (provided through a mechanical ventilation system) and of the percentage of vaccination.

As mentioned in the methodology (section 2.4), for mechanically-ventilated classrooms, the Revent < 1 condition can be maintained if the required AERs obtained for the selected scenarios are adopted. In particular, in that case, a simple constant air volume flow system is enough to provide the required AER and no complex control algorithms, typical of demand-controlled ventilation systems, are needed. In fact, once the scenario is defined, in principle no feedback information is required: a possible procedure in the case of schools equipped with mechanical ventilation is schematically presented in Figure 4. In particular, data regarding the expected scenario (e.g. teacher giving lesson for the first 60 min of the school-day using a microphone; total exposure time; classroom volume) should be provided to the control unit (e.g. inputting through a user input screen) that will be able to evaluate the required AER on the basis of the equations reported in the section 2.1 and, consequently, will set the needed air flow rate of the mechanical ventilation system. In other words, different modes of operation can be selected based on pre-determined activities and durations for the classroom (e.g. lecture, lunch, exercise activity, etc.). Optimizing the provided ventilation based on demand is important given the high energy cost of conditioning outside air in many climates [60–64]; however, our proposed strategy for practical infection control is based on the activities of the occupants rather than CO2 a *priori*. Where activity schedules are consistent, modes of operation can be scheduled in advance to eliminate the need for constant adjustment.

**Figure 4.**
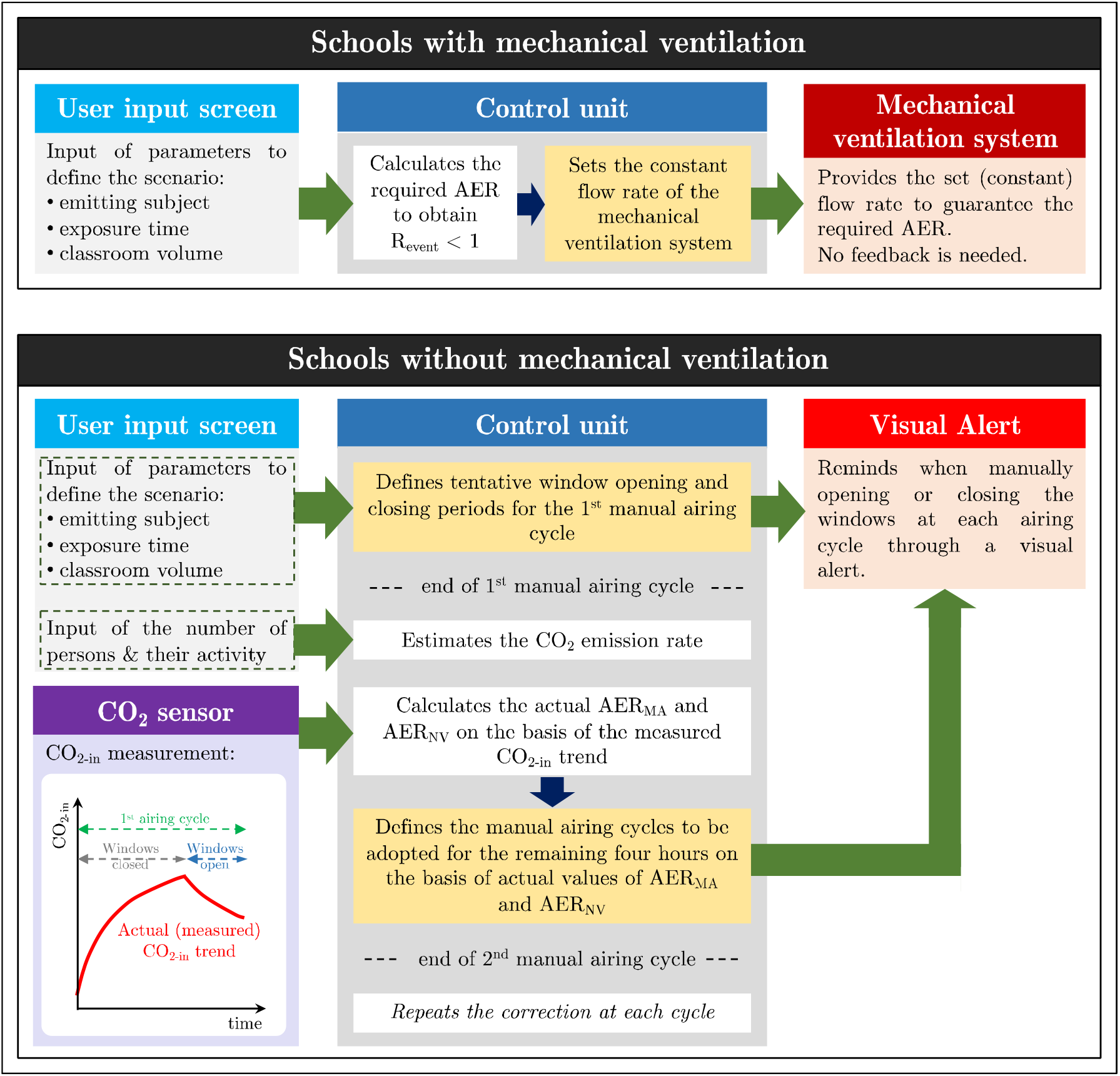
Scheme of the suggested procedures to be applied in schools with and without mechanical ventilation to maintain R_event_ < 1.

### 3.2 Airing procedures for naturally-ventilated schools

In the case of school without mechanical ventilation, maintaining a Revent < 1 is a challenge for scenarios characterized by high emitting infected subjects for two main reasons: i) keeping the windows opened could be not enough to guarantee very high fresh air flow rates, ii) keeping the windows opened for long periods could be detrimental for thermal comfort and energy conservation purposes [58,60,65]. Adopting manual airing cycles described in the section 2.4 represents a practical solution, but it should be kept in mind that the scheduling of window opening and closing period can affect the infection risk of the exposed susceptibles and a required AER cannot be determined *a-priori*. As an example, if AER_NV_ and AER_MA_ were a constant 0.2 and 4.0 h-1, respectively, for the scenario T-60-S, a R_event_ < 1 (i.e. R = 4.2%) could be obtained opening the windows for about 10 min at the end of each hour. The resulting school-day average AER would be equal to of 0.8 h^-1^, which similar to that needed in case of constant mechanical ventilation systems. But, for lower AERs, e.g. constant AER_NV_ and AER_MA_ equal to 0.15 and 2.0 h^-1^, respectively, the required opening period at the end of each hour is 36 min then resulting in a school-day average AER of 1.3 h^-1^ which is significantly higher than that required in the case of steady state mechanical ventilation system. These two easy examples, highlight that the lower the AER_NV_ and AER_MA_ values, the longer the required airing period and, consequently, the higher the resulting school-day average AER.

Thus, the airing strategies are strongly affected by the air exchange rate values, therefore adopting scheduled airing procedures could be misleading. In October 2020 the German Environment Agency (UBA) issued guidance for schools recommending airing classrooms for 5 minutes after every 20 minutes (www.umweltbundesamt.de/en/press/pressinformation/coronavirus-protection-in-schools-airing-rooms-for). For the T-60-S scenario, in the likely hypothesis of having AER_NV_ at least equal to 0.2 h^-1^, adopting the German procedure would allow maintaining a R_event_ < 1 only for AER_MA_ > 3.3 h^-1^; however, such an air exchange rate during manual airing could be not reached.

The above-mentioned examples highlight how AER_NV_ and AER_MA_ need to be continuously monitored so that the airing procedure can be adjusted in real-time. This procedure is obviously more complex than providing mechanical ventilation, and the support of control unit is even more important as it should be able to communicate with a CO_2_ sensor and provide visual alerts on when opening and closing the windows, which will be performed manually by personnel in the classroom (e.g. teacher). Alternatively, since it may be challenging to have a teacher or student reliably open and close a window in response to frequent prompts from the control unit, for relatively minor incremental cost one or more windows in the room could be fitted with a motorized louver, or damper, connected to the control unit such that the percent open of the window can be automatically adjusted by the system. Anyway, whatever the windows are automatically or manually opened, the ventilation procedure is equivalent and provided by the control unit. Indeed, data regarding the expected scenario will be provided to the control unit (through the user input screen as well), then the control unit will evaluate and suggest the manual airing procedure to be adopted in order to guarantee a R_event_ < 1. In particular, the control unit will use as feedback information the indoor CO_2_ concentration continuously measured by an in-room sensor and, on the basis of the number of persons and their activity levels (that will be provided through the user input screen) and of the initial indoor CO_2_ concentration, it will back-calculate the actual AERs during both the period with windows close (AER_NV_) and open (AER_MA_) using the CO_2_ mass balance equation (eq. 7) (Figure 4). Such calculation should be performed adopting a multi-points method, i.e. finding the best regression fit to the continuous CO_2_ data, which is more accurate than the two-points method (i.e. considering just the CO_2_ measurement of start point and end point of natural ventilation and manual airing periods) [66] since it will be less affected by CO_2_ sensor accuracy and intermittent “noisy” measurements. On the basis of the actual AERs the corrected t_MA_ and t_NV_ periods will be calculated by the control unit and the windows opening periods will be scheduled as well for the further four hours (i.e. four cycles) in order to obtain a R_event_ < 1. Since the AER_NV_ and AER_MA_ values are not known *a-priori*, during the first hour/cycle tentative opening and closing periods can be adopted (e.g. 50 min with windows closed and 10 min with windows open). Then, the measurement of the actual AERs will allow scheduling the equally-spaced opening periods of the remaining four hours in order to obtain a R_event_ < 1 (i.e. R = 4.2%) including the entire school-day (i.e. five hours) in the calculation. The scheduled opening and closing periods also consider that if the infected teacher gives lesson on the second, third, fourth or fifth hour the R_event_ < 1 condition must be verified. Actually, the latest the infected teacher enters the classroom, the shorter the exposure period of the susceptible persons (students in this case), this is the reason why we have considered the first hour in the simulations as worst scenario. Counterintuitively, for very different airing periods amongst the airing cycles, the resulting risk for exposed people could be higher for teacher entering the classroom on the last hours than on the first ones, illustrating the need for real-time feedback on what is going on in the classroom. This could be partially solved scheduling equally-spaced opening periods as mentioned above; nonetheless, the control unit needs to check if the opening periods based on actual AERs can guarantee a R_event_ < 1 also for infected teacher entering the classroom at different hours.

At the end of the second cycle AER_NV_ and AER_MA_ will be back-calculated again and, in case, the opening and closing periods will be modified again. Indeed, a high variability of the air exchange rate with windows open could occur, thus, significant corrections may be needed.

An example application of the correction procedure is presented in Figure 5 for the scenario T-60-S. In the figure the indoor CO_2_ concentration, SARS-CoV-2 quanta concentration, and individual risk trend are presented. During the first hour a tentative airing cycle made up of 50 min with windows closed and 10 min with windows open was adopted. From the CO_2_ trend, the actual AER_NV_ and AER_MA_ values were back-calculated and (in this illustrative example) are equal to 0.15 and 2.0 h^-1^, respectively. On the basis of the actual AERs, in order to maintain a R_event_ < 1 (i.e. R = 4.2%), the control unit schedules equally-spaced window opening periods of 42 min for the remaining four hours to be applied at the end of each hour. Thus, the total times during which the windows were kept closed and open for the entire school day are t_NV_ = 122 min and t_MA_ = 178 min (having included the 50 min and 10 min of window closing and opening periods of the first hour) then resulting in a school-day average AER of about 1.3 h^-1^. The tentative opening and closing periods adopted for the first hour were then too short compared to the actual low AERs, for this reason the quanta concentration in the first hour increases significantly and the individual risk trend as well with respect to the same scenario occurring in a classroom equipped with a mechanical ventilation system where a constant AER = 0.8 h^-1^ is enough to maintain R_event_ < 1. The scheduled opening and closing periods also maintain a R_event_ < 1 if the infected teacher gave lesson in the second (R = 4.1%), third (R = 4.1%), fourth (R = 3.8%) or fifth hour (R = 2.4%). In this example the actual AERs were considered constant during the entire school-day, nonetheless, if the AERs at the end of each closing and opening periods do not match with the expected ones (0.15 and 2.0 h^-1^ in this example) further corrections are needed at the end of each hour.

**Figure 5.**
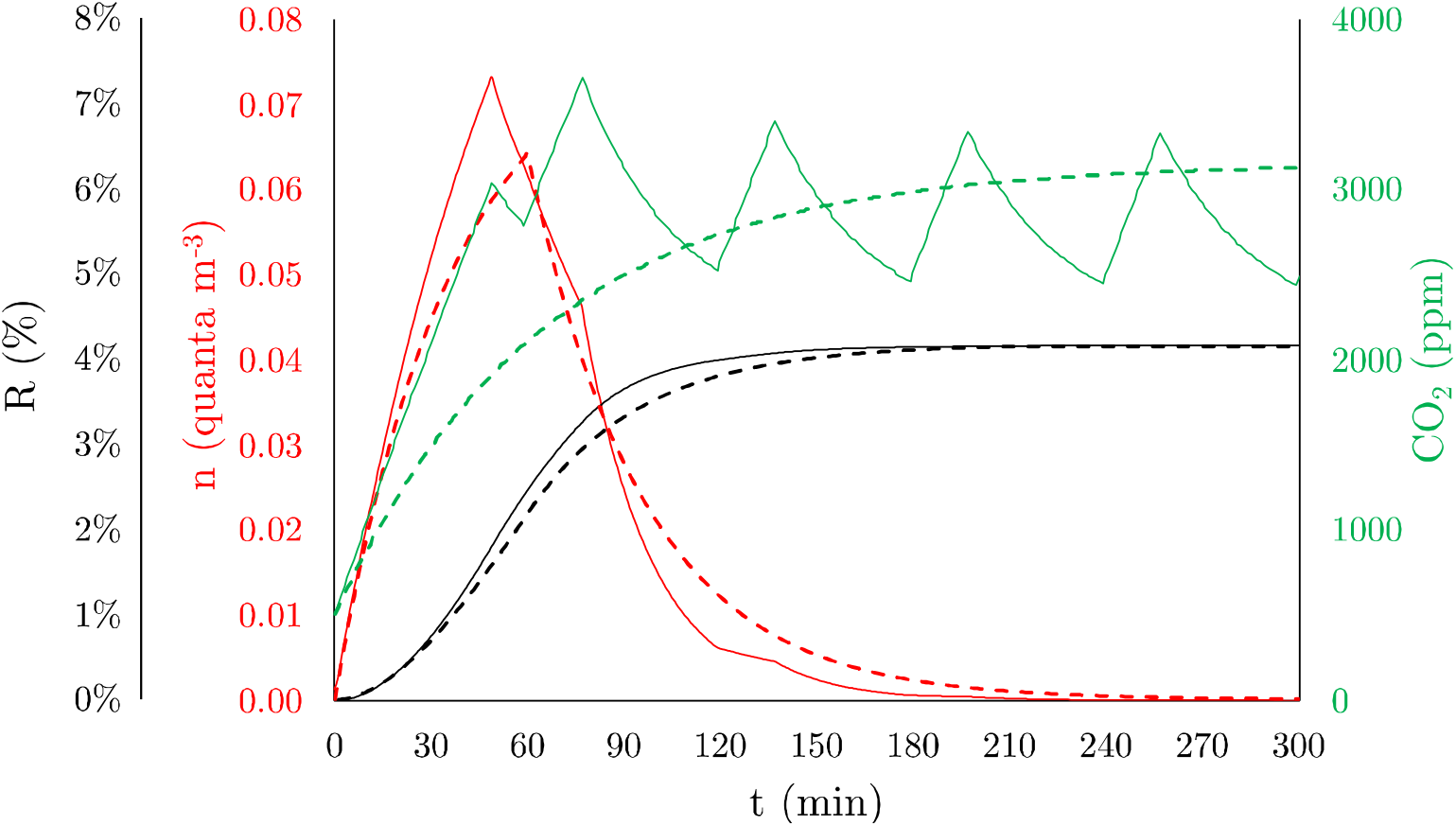
Trends of quanta concentration (n), individual risk (R), and indoor CO_2_ concentration (CO_2-in_) for the scenario T-60-S in the case of SARS-CoV-2 to maintain a R_event_ < 1 through (a) mechanical ventilation system (constant AER = 0.8 h^-1^; bold dotted lines) and (b) manual airing procedures corrected for actual AER (school-day average AER = 1.3 h^-1^ in the hypothesis of measured AER_NV_ and AER_MA_ of 0.15 and 2.0 h^-1^, respectively; thin solid lines).

### 3.3 Applicability and limitation of the methodology for ventilation control

The methodology presented in the paper addresses the proposed goals of (i) quantifying the required ventilation (provided through mechanical systems or manual airing) to reduce the spread of infectious diseases via the airborne route and (ii) proposing a suitable feedback control strategy to monitor and adjust such ventilation in naturally-ventilated classrooms. Nonetheless, in order to effectively reduce the transmission potential of a disease, the uncertainty of the event reproduction number (R_event_) should be taken into account such that, the required air exchange rate maintains (R_event_ -U_Revent_) < 1, with U_Revent_ representing the expanded uncertainty (e.g. with a coverage factor of 95%). The evaluation of U_Revent_ cannot be easily evaluated as it depends on several parameters and models adopted in the calculations presented in the section 2.1 (eq. 1-5). Indeed, when evaluating the R_event_ (eq. 1-5) the following data are needed: quanta emission rate (ER_q_), deposition rate (k), inactivation rate (λ), inhalation rate (IR), room volume (V), air exchange rate (AER), time of exposure (T). The quanta emission rate was investigated in our previous papers where we highlighted that uncertainty relates to the quality of data on viral load, infectious dose and particle volume: such data, at least for SARS-CoV-2, are not definitive [44,51,67] also due to the presence of different viral lineages [68]. Therefore, even if the ER_q_ data provided by our model are much more suitable than those typically estimated based on retrospective assessments of infectious outbreaks, a not negligible uncertainty exists. The deposition rate is mainly affected by the particle size [55] and, thus, adopting an average parameter, as typical of easy-to-use box-models, results in additional uncertainty as well; similarly, data on the virus inactivation rate for SARS-CoV-2 are still limited [53,69]. The inhalation rate depends on the activity levels of the subject; different scientific papers [42,70] reported different IR values for the same activity then confirming a significant variability as well. Room volume and time of exposure can be considered as fixed values (or at least with a not significant uncertainty contribution) as well as the AER if provided through a mechanical ventilation system. The uncertainty budget should also include the physical limitations of the box model (i.e. homogeneous concentration within the room), particle dosimetry model, and dose-response model as well.

The uncertainty budget would be even more complex for confined spaces without mechanical ventilation where manual airing procedures, corrected on the basis of the measured CO_2_ values, are put in place. Indeed, in this case, the uncertainty of the CO_2_ measurements and of the CO_2_ mass balance equation (“constant injection rate method” [32,47]) to back-calculate the corrected AERs should be included too. The effect of the CO_2_ measurement uncertainty is quite straightforward: indeed, in view of correcting the manual airing cycles on the basis of the CO_2_ measurement, a higher CO_2_ uncertainty would undermine the back-calculation of the actual AERs. The CO_2_ measurement uncertainty is typically affected by the sensor accuracy, resolution, temperature effect, static pressure effect, dew-point effect, and probe positioning within the room [71]. CO_2_ probes should be able to provide measurement data with an expanded accuracy of about 5% [71,72], but low-cost sensors may presents larger uncertainties. Nonetheless, as mentioned above, adopting a multi-points method could overcome the problem of the CO_2_ sensor accuracy [66] and significantly reduce the AER uncertainty contribution with respect to two-point method. Indeed, the CO_2_ measurement uncertainty could represent a secondary contribution to the AER back-calculation through the eq. 7, in fact, the uncertainty of the exhaled CO_2_ emission rate could mainly affect the AER uncertainties [73,74].

Summarizing, the uncertainty budget of R_event_ is quite complex and beyond the aims of the current paper. Further studies, in particular those applying real-world measurements and data, are needed in view of improving the quantification of the virus transmission potential for different ventilation systems.

## 4 Conclusions

The study provides a method to support regulatory authorities in the safe operation of schools in the time of pandemics. To this end the required ventilation to reduce the spread of infectious diseases via the airborne route was assessed for both mechanically-and naturally-ventilated classrooms through virus mass balance equations. For the latter, which represent the more frequent and also the more challenging situation, a suitable feedback control strategy based on exhaled CO_2_ monitoring was also proposed in view of maintaining a limited transmission potential of the disease.

The scenarios simulated revealed that adopting a CO_2_ concentration threshold as a possible proxy for virus transmission can be misrepresentative. Indeed, the dynamics of the virus-laden particles and the occurrence of the virus emission may strongly differ from the exhaled CO_2_ ones, thus, CO_2_ and virus concentrations (expressed as “quanta” concentrations) may present significantly different trends. Seasonal influenza presents a negligible transmission potential via airborne route in classroom, even when low ventilation is provided; this is due to the low emission rates typical of such virus. On the contrary, the required air exchange rates to guarantee a R_event_ < 1 for SARS-CoV-2 can be very high for scenarios characterized by highly-emitting infected subjects, such as teacher loudly speaking. Such AERs could be even higher than those suggested by the indoor air quality technical standards, thus, mitigation solutions (e.g. voice modulation in particular) or adequate immunization coverage (i.e. high vaccination percentage) are welcomed.

In order to reduce the virus transmission potential, *ad-hoc* procedures were defined in the case of both mechanically- and naturally-ventilated classrooms. For mechanically-ventilated classrooms a very straightforward procedure was defined since, once the scenario (in terms of emitting subject, classroom geometry, etc.), a control unit can calculate the required air exchange rate accordingly and set the corresponding constant fresh flow rate of the mechanical ventilation system. Such a scenario can be established as a selectable mode of operation for the control unit.

For naturally-ventilated classrooms a suitable feedback control strategy was included and applied in the method. In these classrooms, manual airing cycles help increase the air exchange rate but, due to the dynamic of the emission and of the airing cycles, a required air exchange rate cannot be defined a-*priori*. Thus, the design parameter is not just the air exchange rate but the R_event_ < 1 condition itself which informs scheduling of the manual airing procedures. Such manual airing would be continuously checked and, in case, re-scheduled on the basis of the indoor CO_2_ concentration monitoring. The monitoring would allow evaluation of the actual ventilation rates during the airing cycles and inform proper adjustments to the airing periods.

While further efforts are needed to quantify and reduce the uncertainties of the models, parameters and measured data in the evaluation of individual risk and virus transmission potential, the suggested method provide critical support for national public health authorities to minimize the contribution of school environments to the spread of the pandemics.

## Data Availability

-

## References

[1] S. Chang, E. Pierson, P.W. Koh, J. Gerardin, B. Redbird, D. Grusky, J. Leskovec, Mobility network models of COVID-19 explain inequities and inform reopening., Nature. 589 (2021) 82–87. https://doi.org/10.1038/s41586-020-2923-3.

[2] S.L. Miller, W.W. Nazaroff, J.L. Jimenez, A. Boerstra, G. Buonanno, S.J. Dancer, J. Kurnitski, L.C. Marr, L. Morawska, C. Noakes, Transmission of SARS-CoV-2 by inhalation of respiratory aerosol in the Skagit Valley Chorale superspreading event, Indoor Air. n/a (2020). https://doi.org/10.1111/ina.12751.

[3] L. Morawska, J.W. Tang, W. Bahnfleth, P.M. Bluyssen, A. Boerstra, G. Buonanno, J. Cao, S. Dancer, A. Floto, F. Franchimon, C. Haworth, J. Hogeling, C. Isaxon, J.L. Jimenez, J. Kurnitski, Y. Li, M. Loomans, G. Marks, L.C. Marr, L. Mazzarella, A.K. Melikov, S. Miller, D.K. Milton, W. Nazaroff, P.V. Nielsen, C. Noakes, J. Peccia, X. Querol, C. Sekhar, O. Seppänen, S. Tanabe, R. Tellier, K.W. Tham, P. Wargocki, A. Wierzbicka, M. Yao, How can airborne transmission of COVID-19 indoors be minimised?, Environment International. 142 (2020) 105832. https://doi.org/10.1016/j.envint.2020.105832.

[4] B. Blocken, T. van Druenen, T. van Hooff, P.A. Verstappen, T. Marchal, L.C. Marr, Can indoor sports centers be allowed to re-open during the COVID-19 pandemic based on a certificate of equivalence?, Building and Environment. 180 (2020) 107022. https://doi.org/10.1016/j.buildenv.2020.107022.

[5] G. Buonanno, L. Morawska, L. Stabile, Quantitative assessment of the risk of airborne transmission of SARS-CoV-2 infection: Prospective and retrospective applications, Environment International. 145 (2020) 106112. https://doi.org/10.1016/j.envint.2020.106112.

[6] G. Buonanno, L. Stabile, L. Morawska, Estimation of airborne viral emission: Quanta emission rate of SARS-CoV-2 for infection risk assessment, Environment International. 141 (2020) 105794. https://doi.org/10.1016/j.envint.2020.105794.

[7] T.P. Baggett, H. Keyes, N. Sporn, J.M. Gaeta, Prevalence of SARS-CoV-2 Infection in Residents of a Large Homeless Shelter in Boston, JAMA. 323 (2020) 2191–2192. https://doi.org/10.1001/jama.2020.6887.

[8] Y. Li, G.M. Leung, J.W. Tang, X. Yang, C.Y.H. Chao, J.Z. Lin, J.W. Lu, P.V. Nielsen, J. Niu, H. Qian, A.C. Sleigh, H.-J.J. Su, J. Sundell, T.W. Wong, P.L. Yuen, Role of ventilation in airborne transmission of infectious agents in the built environment - a multidisciplinary systematic review., Indoor Air. 17 (2007) 2–18. https://doi.org/10.1111/j.1600-0668.2006.00445.x.

[9] R.M. Viner, S.J. Russell, H. Croker, J. Packer, J. Ward, C. Stansfield, O. Mytton, C. Bonell, R. Booy, School closure and management practices during coronavirus outbreaks including COVID-19: a rapid systematic review, School Closure and Management Practices during Coronavirus Outbreaks Including COVID-19: A Rapid Systematic Review. (2020). https://doi.org/10.1016/s2352-4642(20)30095-x.

[10] M. Klimek-Tulwin, T. Tulwin, Early school closures can reduce the first-wave of the COVID-19 pandemic development, Journal of Public Health. (2020). https://doi.org/10.1007/s10389-020-01391-z.

[11] J. Bayham, E.P. Fenichel, Impact of school closures for COVID-19 on the US health-care workforce and net mortality: a modelling study, The Lancet Public Health. 5 (2020) e271–e278. https://doi.org/10.1016/S2468-2667(20)30082-7.

[12] J. Zhang, Y. Hayashi, L.D. Frank, COVID-19 and transport: Findings from a world-wide expert survey, Transport Policy. 103 (2021) 68–85. https://doi.org/10.1016/j.tranpol.2021.01.011.

[13] K. Farsalinos, K. Poulas, D. Kouretas, A. Vantarakis, M. Leotsinidis, D. Kouvelas, A.O. Docea, R. Kostoff, G.T. Gerotziafas, M.N. Antoniou, R. Polosa, A. Barbouni, V. Yiakoumaki, T.V. Giannouchos, P.G. Bagos, G. Lazopoulos, B.N. Izotov, V.A. Tutelyan, M. Aschner, T. Hartung, H.M. Wallace, F. Carvalho, J.L. Domingo, A. Tsatsakis, Improved strategies to counter the COVID-19 pandemic: Lockdowns vs. primary and community healthcare, Toxicology Reports. 8 (2021) 1–9. https://doi.org/10.1016/j.toxrep.2020.12.001.

[14] D.R. Petretto, I. Masala, C. Masala, School Closure and Children in the Outbreak of COVID-19, Clin Pract Epidemiol Ment Health. 16 (2020) 189–191. https://doi.org/10.2174/1745017902016010189.

[15] W. Chen, N. Zhang, J. Wei, H.-L. Yen, Y. Li, Short-range airborne route dominates exposure of respiratory infection during close contact, Building and Environment. 176 (2020) 106859. https://doi.org/10.1016/j.buildenv.2020.106859.

[16] Z.T. Ai, A.K. Melikov, Airborne spread of expiratory droplet nuclei between the occupants of indoor environments: A review., Indoor Air. 28 (2018) 500–524. https://doi.org/10.1111/ina.12465.

[17] Z.T. Ai, K. Hashimoto, A.K. Melikov, Airborne transmission between room occupants during short-term events: Measurement and evaluation, Indoor Air. 29 (2019) 563–576. https://doi.org/10.1111/ina.12557.

[18] W.J. Edmunds, Finding a path to reopen schools during the COVID-19 pandemic, The Lancet Child & Adolescent Health. 4 (2020) 796–797. https://doi.org/10.1016/S2352-4642(20)30249-2.

[19] N. Ziauddeen, K. Woods-Townsend, S. Saxena, R. Gilbert, N.A. Alwan, Schools and COVID-19: Reopening Pandora’s box?, Public Health in Practice. 1 (2020) 100039. https://doi.org/10.1016/j.puhip.2020.100039.

[20] N.H.L. Leung, D.K.W. Chu, E.Y.C. Shiu, K.-H. Chan, J.J. McDevitt, B.J.P. Hau, H.-L. Yen, Y. Li, D.K.M. Ip, J.S.M. Peiris, W.-H. Seto, G.M. Leung, D.K. Milton, B.J. Cowling, Respiratory virus shedding in exhaled breath and efficacy of face masks, Nature Medicine. 26 (2020) 676–680. https://doi.org/10.1038/s41591-020-0843-2.

[21] L. Morawska, J. Cao, Airborne transmission of SARS-CoV-2: The world should face the reality, Environment International. 139 (2020) 105730. https://doi.org/10.1016/j.envint.2020.105730.

[22] R. Tellier, Aerosol transmission of influenza A virus: a review of new studies., J R Soc Interface. 6 Suppl 6 (2009) S783–790. https://doi.org/10.1098/rsif.2009.0302.focus.

[23] S. Tang, Y. Mao, R.M. Jones, Q. Tan, J.S. Ji, N. Li, J. Shen, Y. Lv, L. Pan, P. Ding, X. Wang, Y. Wang, C.R. MacIntyre, X. Shi, Aerosol transmission of SARS-CoV-2? Evidence, prevention and control, Environ Int. 144 (2020) 106039–106039. https://doi.org/10.1016/j.envint.2020.106039.

[24] S.N. Rudnick, D.K. Milton, Risk of indoor airborne infection transmission estimated from carbon dioxide concentration., Indoor Air. 13 (2003) 237–245. https://doi.org/10.1034/j.1600-0668.2003.00189.x.

[25] P. de Man, S. Paltansing, D.S.Y. Ong, N. Vaessen, G. van Nielen, J.G.M. Koeleman, Outbreak of Coronavirus Disease 2019 (COVID-19) in a Nursing Home Associated With Aerosol Transmission as a Result of Inadequate Ventilation, Clinical Infectious Diseases. (2020). https://doi.org/10.1093/cid/ciaa1270.

[26] R.M. Baloch, C.N. Maesano, J. Christoffersen, S. Banerjee, M. Gabriel, É. Csobod, E. de Oliveira Fernandes, I. Annesi-Maesano, É. Csobod, P. Szuppinger, R. Prokai, P. Farkas, C. Fuzi, E. Cani, J. Draganic, E.R. Mogyorosy, Z. Korac, E. de Oliveira Fernandes, G. Ventura, J. Madureira, I. Paciência, A. Martins, R. Pereira, E. Ramos, P. Rudnai, A. Páldy, G. Dura, T. Beregszászi, É. Vaskövi, D. Magyar, T. Pándics, Z. Remény-Nagy, R. Szentmihályi, O. Udvardy, M.J. Varró, S. Kephalopoulos, D. Kotzias, J. Barrero-Moreno, R. Mehmeti, A. Vilic, D. Maestro, H. Moshammer, G. Strasser, P. Brigitte, P. Hohenblum, E. Goelen, M. Stranger, M. Spruy, M. Sidjimov, A. Hadjipanayis, A. Katsonouri-Sazeides, E. Demetriou, R. Kubinova, H. Kazmarová, B. Dlouha, B. Kotlík, H. Vabar, J. Ruut, M. Metus, K. Rand, A. Järviste, A. Nevalainen, A. Hyvarinen, M. Täubel, K. Järvi, I. Annesi-Maesano, C. Mandin, B. Berthineau, H.-J. Moriske, M. Giacomini, A. Neumann, J. Bartzis, K. Kalimeri, D. Saraga, M. Santamouris, M.N. Assimakopoulos, V. Asimakopoulos, P. Carrer, A. Cattaneo, S. Pulvirenti, F. Vercelli, F. Strangi, E. Omeri, S. Piazza, A. D’Alcamo, A.C. Fanetti, P. Sestini, M. Kouri, G. Viegi, G. Sarno, S. Baldacci, S. Maio, S. Cerrai, V. Franzitta, S. Bucchieri, F. Cibella, M. Simoni, M. Neri, D. Martuzevičius, E. Krugly, S. Montefort, P. Fsadni, P.Z. Brewczyński, E. Krakowiak, J. Kurek, E. Kubarek, A. Wlazło, C. Borrego, C. Alves, J. Valente, E. Gurzau, C. Rosu, G. Popita, I. Neamtiu, C. Neagu, D. Norback, P. Bluyssen, M. Bohms, P. Van Den Hazel, F. Cassee, Y.B. de Bruin, A. Bartonova, A. Yang, K. Halzlová, M. Jajcaj, M. Kániková, O. Miklankova, M. Vítkivá, M. Jovsevic-Stojanovic, M. Zivkovic, Z. Stevanovic, I. Lazovic, Z. Stevanovic, Z. Zivkovic, S. Cerovic, J. Jocic-Stojanovic, D. Mumovic, P. Tarttelin, L. Chatzidiakou, E. Chatzidiakou, M.-C. Dewolf, Indoor air pollution, physical and comfort parameters related to schoolchildren’s health: Data from the European SINPHONIE study, Science of The Total Environment. 739 (2020) 139870. https://doi.org/10.1016/j.scitotenv.2020.139870.

[27] E. Csobod Annesi-Maesano, I Carrer, P.,. Kephalopoulos, S.,. Madureira, J.,. Rudnai, P.,. De Oliveira Fernandes, E.,. Barrero, J.,. Beregszászi, T.,. Hyvärinen, A.,. Moshammer, H.,. Norback, D.,. Páldy, A.,. Pándics, T.,. Sestini, P.,. Stranger, M.,. Taubel, M.,. Varró, M.,. Vaskovi, E.,. Ventura Silva, G. Viegi, G.,., SINPHONIE – Schools Indoor Pollution and Health Observatory Network in Europe - Final Report, Publications Office of the European Union, Luxembourg, 2014.

[28] S. Zhu, S. Jenkins, K. Addo, M. Heidarinejad, S.A. Romo, A. Layne, J. Ehizibolo, D. Dalgo, N.W. Mattise, F. Hong, O.O. Adenaiye, J.P. Bueno de Mesquita, B.J. Albert, R. Washington-Lewis, J. German, S. Tai, S. Youssefi, D.K. Milton, J. Srebric, Ventilation and laboratory confirmed acute respiratory infection (ARI) rates in college residence halls in College Park, Maryland, Environment International. 137 (2020) 105537. https://doi.org/10.1016/j.envint.2020.105537.

[29] B. Pavilonis, A.M. Ierardi, L. Levine, F. Mirer, E.A. Kelvin, Estimating Aerosol Transmission Risk of SARS-CoV-2 in New York City Public Schools During Reopening, Environ Res. (2021) 110805– 110805. https://doi.org/10.1016/j.envres.2021.110805.

[30] A. Pacitto, L. Stabile, L. Morawska, M. Nyarku, M.A. Torkmahalleh, Z. Akhmetvaliyeva, A. Andrade, F.H. Dominski, P. Mantecca, W.H. Shetaya, M. Mazaheri, R. Jayaratne, S. Marchetti, S.K. Hassan, A. El-Mekawy, E.F. Mohamed, L. Canale, A. Frattolillo, G. Buonanno, Daily submicron particle doses received by populations living in different low- and middle-income countries, Environmental Pollution. 269 (2021) 116229. https://doi.org/10.1016/j.envpol.2020.116229.

[31] L. Stabile, G. Buonanno, A. Frattolillo, M. Dell’Isola, The effect of the ventilation retrofit in a school on CO2, airborne particles, and energy consumptions, Building and Environment. 156 (2019) 1–11. https://doi.org/10.1016/j.buildenv.2019.04.001.

[32] N. Mahyuddin, H.B. Awbi, A Review of CO2 Measurement Procedures in Ventilation Research, International Journal of Ventilation. 10 (2012) 353–370. https://doi.org/10.5555/2044-4044-10.4.353.

[33] Z. Bakó-Biró, D.J. Clements-Croome, N. Kochhar, H.B. Awbi, M.J. Williams, Ventilation rates in schools and pupils’ performance, Building and Environment. 48 (2) 215–223. https://doi.org/10.1016/j.buildenv.2011.08.018.

[34] M.J. Mendell, E.A. Eliseeva, M.M. Davies, M. Spears, A. Lobscheid, W.J. Fisk, M.G. Apte, Association of classroom ventilation with reduced illness absence: a prospective study in California elementary schools., Indoor Air. 23 (2013) 515–528. https://doi.org/10.1111/ina.12042.

[35] C. Zemouri, S.F. Awad, C.M.C. Volgenant, W. Crielaard, A.M.G.A. Laheij, J.J. de Soet, Modeling of the Transmission of Coronaviruses, Measles Virus, Influenza Virus, Mycobacterium tuberculosis, and Legionella pneumophila in Dental Clinics., J Dent Res. 99 (2020) 1192–1198. https://doi.org/10.1177/0022034520940288.

[36] R.K. Bhagat, M.S. Davies Wykes, S.B. Dalziel, P.F. Linden, Effects of ventilation on the indoor spread of COVID-19, Journal of Fluid Mechanics. 903 (2020) F1. https://doi.org/10.1017/jfm.2020.720.

[37] S.-Y. Cheng, C.J. Wang, A.C.-T. Shen, S.-C. Chang, How to Safely Reopen Colleges and Universities During COVID-19: Experiences From Taiwan, Ann Intern Med. 173 (2020) 638–641. https://doi.org/10.7326/M20-2927.

[38] A. Mikszewski, L. Stabile, G. Buonanno, L. Morawska, THE AIRBORNE CONTAGIOUSNESS OF RESPIRATORY VIRUSES: A COMPARATIVE ANALYSIS AND IMPLICATIONS FOR MITIGATION, MedRxiv. (2021) 2021.01.26.21250580. https://doi.org/10.1101/2021.01.26.21250580.

[39] P. Tupper, H. Boury, M. Yerlanov, C. Colijn, Event-specific interventions to minimize COVID-19 transmission, Proc Natl Acad Sci USA. 117 (2020) 32038. https://doi.org/10.1073/pnas.2019324117.

[40] T. Moreno, R.M. Pintó, A. Bosch, N. Moreno, A. Alastuey, M.C. Minguillón, E. Anfruns-Estrada, S. Guix, C. Fuentes, G. Buonanno, L. Stabile, L. Morawska, X. Querol, Tracing surface and airborne SARS-CoV-2 RNA inside public buses and subway trains, Environment International. 147 (2021) 106326. https://doi.org/10.1016/j.envint.2020.106326.

[41] B.G. Wagner, B.J. Coburn, S. Blower, Calculating the potential for within-flight transmission of influenza A (H1N1), BMC Medicine. 7 (2009) 81. https://doi.org/10.1186/1741-7015-7-81.

[42] W.C. Adams, Measurement of Breathing Rate and Volume in Routinely Performed Daily Activities. Final Report. Human Performance Laboratory, Physical Education Department, University of California, Davis., Human Performance Laboratory, Physical Education Department, University of California, Davis. Prepared for the California Air Resources Board, Contract No. A033-205, April 1993., 1993.

[43] International Commission on Radiological Protection, Human respiratory tract model for radiological protection. A report of a Task Group of the International Commission on Radiological Protection., Annals of the ICRP. 24 (1994) 1–482. https://doi.org/10.1016/0146-6453(94)90029-9.

[44] T. Watanabe, T.A. Bartrand, M.H. Weir, T. Omura, C.N. Haas, Development of a dose-response model for SARS coronavirus., Risk Anal. 30 (2010) 1129–1138. https://doi.org/10.1111/j.1539-6924.2010.01427.x.

[45] G.N. Sze To, C.Y.H. Chao, Review and comparison between the Wells–Riley and dose-response approaches to risk assessment of infectious respiratory diseases, Indoor Air. 20 (2010) 2–16. https://doi.org/10.1111/j.1600-0668.2009.00621.x.

[46] A. Persily, L. de Jonge, Carbon dioxide generation rates for building occupants, Indoor Air. 27 (2017) 868–879. https://doi.org/10.1111/ina.12383.

[47] W.W. Nazaroff, Residential air-change rates: A critical review, Indoor Air. n/a (2021). https://doi.org/10.1111/ina.12785.

[48] European Committee for Standardisation, UNI EN 15251 - Indoor environmental input parameters for design and assessment of energy performance of buildings addressing indoor air quality, thermal environment, lighting and acoustics, (2008).

[49] R.H. Alford, J.A. Kasel, P.J. Gerone, V. Knight, Human influenza resulting from aerosol inhalation., Proc Soc Exp Biol Med. 122 (1966) 800–804. https://doi.org/10.3181/00379727-122-31255.

[50] P.J. Bueno de Mesquita, C.J. Noakes, D.K. Milton, Quantitative aerobiologic analysis of an influenza human challenge-transmission trial, Indoor Air. 30 (2020) 1189–1198. https://doi.org/10.1111/ina.12701.

[51] P. Gale, Thermodynamic equilibrium dose-response models for MERS-CoV infection reveal a potential protective role of human lung mucus but not for SARS-CoV-2, Microb Risk Anal. 16 (2020) 100140–100140. https://doi.org/10.1016/j.mran.2020.100140.

[52] S.E. Eikenberry, M. Mancuso, E. Iboi, T. Phan, K. Eikenberry, Y. Kuang, E. Kostelich, A.B. Gumel, To mask or not to mask: Modeling the potential for face mask use by the general public to curtail the COVID-19 pandemic, Infectious Disease Modelling. 5 (2020) 293–308. https://doi.org/10.1016/j.idm.2020.04.001.

[53] N. van Doremalen, T. Bushmaker, D.H. Morris, M.G. Holbrook, A. Gamble, B.N. Williamson, A. Tamin, J.L. Harcourt, N.J. Thornburg, S.I. Gerber, J.O. Lloyd-Smith, E. de Wit, V.J. Munster, Aerosol and Surface Stability of SARS-CoV-2 as Compared with SARS-CoV-1, N Engl J Med. 382 (2020) 1564–1567. https://doi.org/10.1056/NEJMc2004973.

[54] W. Yang, L.C. Marr, Dynamics of Airborne Influenza A Viruses Indoors and Dependence on Humidity, PLOS ONE. 6 (2011) e21481. https://doi.org/10.1371/journal.pone.0021481.

[55] S.E. Chatoutsidou, M. Lazaridis, Assessment of the impact of particulate dry deposition on soiling of indoor cultural heritage objects found in churches and museums/libraries, Journal of Cultural Heritage. 39 (2019) 221–228. https://doi.org/10.1016/j.culher.2019.02.017.

[56] L. Stabile, M. Dell’Isola, A. Russi, A. Massimo, G. Buonanno, The effect of natural ventilation strategy on indoor air quality in schools, Science of the Total Environment. 595 (2017) 894–902. https://doi.org/10.1016/j.scitotenv.2017.02.030.

[57] F.R. d’Ambrosio Alfano, M. Dell’Isola, G. Ficco, F. Tassini, Experimental analysis of air tightness in Mediterranean buildings using the fan pressurization method, Building and Environment. 53 (2012) 16–25. https://doi.org/10.1016/j.buildenv.2011.12.017.

[58] L. Stabile, M. Dell’Isola, A. Frattolillo, A. Massimo, A. Russi, Effect of natural ventilation and manual airing on indoor air quality in naturally ventilated Italian classrooms, Building and Environment. 98 (2016) 180–189. https://doi.org/10.1016/j.buildenv.2016.01.009.

[59] C. Howard-Reed, L.A. Wallace, W.R. Ott, The Effect of Opening Windows on Air Change Rates in Two Homes, Journal of the Air & Waste Management Association. 52 (2002) 147–159. https://doi.org/10.1080/10473289.2002.10470775.

[60] L. Stabile, A. Massimo, L. Canale, A. Russi, A. Andrade, M. Dell’Isola, The Effect of Ventilation Strategies on Indoor Air Quality and Energy Consumptions in Classrooms, Buildings. 9 (2019). https://doi.org/10.3390/buildings9050110.

[61] L. Canale, M. Dell’Isola, G. Ficco, B. Di Pietra, A. Frattolillo, Estimating the impact of heat accounting on Italian residential energy consumption in different scenarios, Energy and Buildings. 168 (2018) 385–398. https://doi.org/10.1016/j.enbuild.2018.03.040.

[62] M. Dell’Isola, G. Ficco, F. Arpino, G. Cortellessa, L. Canale, A novel model for the evaluation of heat accounting systems reliability in residential buildings, Energy and Buildings. 150 (2017) 281– 293. https://doi.org/10.1016/j.enbuild.2017.06.007.

[63] M. Dell’Isola, G. Ficco, L. Canale, B.I. Palella, G. Puglisi, An IoT Integrated Tool to Enhance User Awareness on Energy Consumption in Residential Buildings, Atmosphere. 10 (2019). https://doi.org/10.3390/atmos10120743.

[64] L. Canale, M. Dell’Isola, G. Ficco, T. Cholewa, S. Siggelsten, I. Balen, A comprehensive review on heat accounting and cost allocation in residential buildings in EU, Energy and Buildings. 202 (2019) 109398. https://doi.org/10.1016/j.enbuild.2019.109398.

[65] A. Heebøll, P. Wargocki, J. Toftum, Window and door opening behavior, carbon dioxide concentration, temperature, and energy use during the heating season in classrooms with different ventilation retrofits—ASHRAE RP1624, Science and Technology for the Built Environment. 24 (2018) 626–637. https://doi.org/10.1080/23744731.2018.1432938.

[66] S. Cui, M. Cohen, P. Stabat, D. Marchio, CO2 tracer gas concentration decay method for measuring air change rate, Building and Environment. 84 (2015) 162–169. https://doi.org/10.1016/j.buildenv.2014.11.007.

[67] M. Abbas, D. Pittet, Surfing the COVID-19 scientific wave, Lancet Infect Dis. (2020) S1473-3099(20)30558–2. https://doi.org/10.1016/S1473-3099(20)30558-2.

[68] C. Alteri, V. Cento, A. Piralla, V. Costabile, M. Tallarita, L. Colagrossi, S. Renica, F. Giardina, F. Novazzi, S. Gaiarsa, E. Matarazzo, M. Antonello, C. Vismara, R. Fumagalli, O.M. Epis, M. Puoti, C.F. Perno, F. Baldanti, Genomic epidemiology of SARS-CoV-2 reveals multiple lineages and early spread of SARS-CoV-2 infections in Lombardy, Italy, Nature Communications. 12 (2021) 434. https://doi.org/10.1038/s41467-020-20688-x.

[69] A.C. Fears, W.B. Klimstra, P. Duprex, A. Hartman, S.C. Weaver, K.C. Plante, D. Mirchandani, J.A. Plante, P.V. Aguilar, D. Fernández, A. Nalca, A. Totura, D. Dyer, B. Kearney, M. Lackemeyer, J.K. Bohannon, R. Johnson, R.F. Garry, D.S. Reed, C.J. Roy, Comparative dynamic aerosol efficiencies of three emergent coronaviruses and the unusual persistence of SARS-CoV-2 in aerosol suspensions, MedRxiv. (2020) 2020.04.13.20063784. https://doi.org/10.1101/2020.04.13.20063784.

[70] D.W. Layton, Metabolically consistent breathing rates for use in dose assessments., Health Phys. 64 (1993) 23–36. https://doi.org/10.1097/00004032-199301000-00003.

[71] L.B. Mendes, N.W.M. Ogink, N. Edouard, H.J.C. van Dooren, I. de F.F. Tinôco, J. Mosquera, NDIR Gas Sensor for Spatial Monitoring of Carbon Dioxide Concentrations in Naturally Ventilated Livestock Buildings, Sensors (Basel). 15 (2015) 11239–11257. https://doi.org/10.3390/s150511239.

[72] M.H. Sherman, I.S. Walker, M.M. Lunden, Uncertainties in Air Exchange using Continuous-Injection, Long-Term Sampling Tracer-Gas Methods, Null. 13 (2014) 13–28. https://doi.org/10.1080/14733315.2014.11684034.

[73] G. Remion, B. Moujalled, M. El Mankibi, Review of tracer gas-based methods for the characterization of natural ventilation performance: Comparative analysis of their accuracy, Building and Environment. 160 (2019) 106180. https://doi.org/10.1016/j.buildenv.2019.106180.

[74] A. Kabirikopaei, J. Lau, Uncertainty analysis of various CO2-Based tracer-gas methods for estimating seasonal ventilation rates in classrooms with different mechanical systems, Building and Environment. 179 (2020) 107003. https://doi.org/10.1016/j.buildenv.2020.107003.

